# Genetic regulation of the plasma proteome and its link to cardiometabolic disease in Greenlandic Inuit

**DOI:** 10.1101/2024.07.03.24309577

**Authors:** Sara E. Stinson, Renzo F. Balboa, Mette K. Andersen, Frederik F. Stæger, Shixu He, Anne Cathrine Baun Thuesen, Long Lin, Peter Bjerregaard, Christina V.L. Larsen, Niels Grarup, Marit E. Jørgensen, Ida Moltke, Anders Albrechtsen, Torben Hansen

**Affiliations:** Novo Nordisk Foundation Center for Basic Metabolic Research, Faculty of Health and Medical Sciences, University of Copenhagen, Copenhagen, Denmark; Department of Biology, University of Copenhagen, Copenhagen, Denmark; National Institute of Public Health, University of Southern Denmark, Copenhagen, Denmark; Greenland Center for Health Research, University of Greenland, Nuuk, Greenland; Steno Diabetes Center, Nuuk, Greenland

**Keywords:** Proteomics, Genomics, Diverse populations, Greenland Inuit, Cardiovascular disease, Type 2 diabetes, pQTL analysis

## Abstract

**Background:** Circulating proteins play essential roles in numerous complex diseases, yet our understanding of the genetic influences on inflammation and cardiovascular-related proteins in non-European isolated populations remains sparse. Here, we aimed to characterise the genetic architecture of plasma protein biomarkers in the Greenlandic population.

**Methods:** Using combined data from Greenlandic population cohorts (n=3,707 individuals), including genotypes and plasma proteomics (177 proteins) from Olink Target 96 Inflammation and Cardiovascular II panels, we performed a protein quantitative trait loci (pQTL) study using a linear mixed model, accounting for relatedness and population structure.

**Findings:** Mapping of 177 plasma proteins in 3,707 adult Greenlandic individuals (mean age 47.9; 54.5% female) reveal 251 primary pQTLs, 235 additive (92 *cis* and 143 *trans*) and 16 recessive (1 *cis* and 15 *trans*), 48 secondary pQTLs, and 70 novel pQTLs (28%). We demonstrate a higher proportion of variance in protein levels explained in Greenlanders compared to Europeans from the UK Biobank (e.g. IL-27, IgGFcRII-b, IL-16, and Gal-9). We describe changes in expression of inflammation and cardiovascular-related proteins associated with known high impact Arctic-specific variants, including in *CPT1A*, *TBC1D4*, *HNF1A*, *LDLR*, and *PCSK9*.

**Interpretation:** These findings highlight the importance of genome-wide plasma proteomic analyses in Greenlanders, and diverse populations in general, with implications for biomarker and therapeutic target development.

**Funding:** Novo Nordisk Foundation, The Independent Research Fund Denmark, and Karen Elise Jensen Foundation.

**Research in context:** *Evidence before this study:* Recent affinity-based proteomic studies have been performed in large European biobank-scale cohorts such as the UK Biobank and deCODE. Several smaller-scale studies have also been performed in isolated European populations, e.g. MANOLIS and Pomak (Hellenic), Orkney (Scotland), and Vis (Croatia). Studies in non-European populations are also beginning to emerge, including in the China Kadoorie Biobank. Studies performed in diverse populations can identify population-specific variants in genes implicated in regulating the expression of proteins, which may be causally linked to inflammation and cardiovascular disease. In particular, small and historically isolated populations, such as the Greenlandic population, are more likely to harbour common variants with larger effect sizes that may contribute to health and disease.

*Added value of this study:* This study reports 251 primary protein quantitative trait loci (pQTLs) associated with the abundance of 177 plasma proteins, 28% of which have not been previously reported. We identified 48 additional pQTLs in a secondary conditional analysis. We identified novel pQTLs that were common in Greenland, but rare globally (e.g. ST1A1, DCN). We found common pQTLs which explained a substantial proportion of variance (>30%) in protein abundance (e.g. IL-27, IgGFcRIIb, IL-16, Gal-9) when compared to Europeans. We examined differences in protein abundance in carriers of Arctic-specific variants (e.g. *CPT1A, TBC1D4*, *HNF1A*, *LDLR*, *PCSK9*) which are implicated in lipid metabolism and cardiometabolic disease, revealing underlying biological mechanisms.

*Implications of all the available evidence:* Given that both genetics and the environment affect protein levels causally linked to disease, it is crucial to perform genome-wide association studies in smaller populations of diverse genetic ancestry to ensure equity in genetic discovery. Investigating the effect of previously identified Arctic-specific variants on protein expression revealed links to therapeutic targets for metabolic disease, which may have implications for the health care system in Greenland and beyond, including access to treatment.

## Introduction

Circulating proteins measured in the blood represent intermediate phenotypes for disease states and are strongly linked to both causative genetic variants and disease outcomes^1^. Thus, differences in protein expression make ideal candidates to study, in order to understand the molecular mechanisms of complex disease.

Proteomics data has only recently become accessible to large-scale genome-wide association studies (GWAS), largely due to analytical limitations specific to the composition of the plasma proteome^2^. Advances in high-throughput proteomics methods (e.g. Olink, SomaScan, and mass-spectrometry approaches) now enable the measurement of plasma levels of hundreds to thousands of proteins concurrently^3,4^. Unlike mass-spectrometry-based proteomics, the Olink Proximity Extension Assay captures less abundant proteins, allowing for the quantification of cytokines and chemokines relevant to the pathophysiology of cardiometabolic disease^5^.

Identification of genetic variants associated with altered plasma protein levels (protein quantitative trait loci, pQTLs) can enhance our understanding of the role of proteins in health and disease^6^. Discovery of genetic determinants of protein levels help elucidate the causal role of a given protein in disease risk, which may allow for improved risk stratification to enhance early diagnosis and identification of potential drug targets for treatment or prevention of such diseases^7^.

To date, the vast majority of studies have restricted analyses of the genetic architecture of the plasma proteome to European ancestries^8–13^, this includes large-scale biobanks such as the UK Biobank Pharma Proteomics Project (UKB-PPP, Olink)^14^ and deCODE Genetics (SomaScan). For example, the discovery analysis in UKB-PPP was limited to European individuals, with replication in a limited number of non-European individuals^14^. Although separate efforts have been made to analyse pQTLs in non-EUR, including East Asian^15,16^ and Middle Eastern^17^ ancestries, diverse populations are in comparison, highly underrepresented.

Two studies have been performed in isolated European populations, including MANOLIS, Pomark, and ORCADES in combination with whole-genome sequencing^18,19^. Studying isolated populations has the power to enhance the discovery of disease-associated loci due to the enrichment of certain rare variants with larger effects^20,21^.

The Greenlandic population, comprising ∼57,000 individuals, mainly has Inuit ancestry, but due to recent admixture it also has some European ancestry^22^. The population is a small and historically isolated founder population, which has experienced an increase in the prevalence of type 2 diabetes (T2D) over the last 25 years^23^. Previous studies of the population have revealed that complex diseases, including T2D, often manifest differently and have Arctic-specific causes, when compared to other global populations. How lipid-related and cardiometabolic disease-causing variants are associated with differences in protein expression within the Greenlandic population, and its comparison with other populations, have yet to be studied. An investigation into these relationships will allow for a better understanding of the mechanism of disease within this population, and other diverse populations in general.

Here, we examined the genetic architecture of the targeted plasma proteome in 3,707 Greenlanders. The aims of this study were to: (i) perform GWAS of 177 plasma proteins measured by the Olink Target 96 Inflammation and Cardiovascular II panels; (ii) compare the genetic architecture of the targeted proteome between Greenlanders and European populations through comparisons with the UKB-PPP, and (iii) assess the impact of known high impact Arctic-specific variants on circulating protein levels.

## Methods

### Ethics statement

The study has received ethical approval from the Science Ethics Committee in Greenland (project 505-42, project 505-95, project 2011–13 [ref. no. 2011–056978], project 2017-5582, project 2015–22 [ref. no. 2015-16426], and project 2021-09 [16669546]). The study was conducted in accordance with the Declaration of Helsinki, second revision. All participants gave written informed consent.

### Study populations

A total of 3,707 participants with available proteomics, genomics, and phenotype data, were included in this study; the participants were pooled from three population-survey cohorts: the Greenland Population Study (B99) (n = 15; 1998–2001)^24^, the Inuit Health in Transition (IHIT) cohort (n = 2,130; 2005–2010)^25^, and the B2018 cohort (n = 1,587; 2017-2019)^26^.

### Plasma proteomics

Relative levels of 184 plasma proteins were measured in 3,707 participants using the Olink Target 96 Inflammation and Cardiovascular II panels across two batches. Seven proteins were common to both panels, yielding a total of 177 unique proteins that were measured. The Olink batches were bridged and normalised using 16 controls using the *OlinkAnalyze* R package (https://cran.r-project.org/web/packages/OlinkAnalyze/index.html). Normalised protein expression values (NPX) on a log_2_ scale were used for downstream analyses. Protein values below the lower limit of detection were included in analyses, while those failing Olink’s internal quality control warnings were excluded.

### Genotyping and imputation

All samples were genotyped using the Multi-Ethnic Genotyping Array (MEGA chip, Illumina), composed of ∼2M variants across the human genome. This chip was used due to the inclusion of variants selected from Indigenous Americans and East Asians, maximising variant capture in Inuit. Genotypes were jointly called using GenCall within GenomeStudio (Illumina). Duplicate samples, individuals with >0.05 missing genotypes, and variants with >0.01 missing genotypes were excluded from analyses. After quality control, our variant set comprised 1.6M variants used for the present study.

The same reference panel described in Senftleber et al.^27^ was utilised in this study. Briefly, whole genome sequencing data from 447 Greenlanders were genotyped using GATK, and phased using SHAPEIT2^28^, using available duo/trio information. These data were then combined with 1000 Genomes^29^ data and utilised as a reference for imputing common (MAF >0.5%) variants using IMPUTE2^30^. The imputed data was used to estimate a genetic relationship matrix (GRM) and admixture proportions for inclusion in genome-wide association analyses, described below.

### Genome-wide association analyses

For association analyses, a linear mixed model (LMM) was used, accounting for relatedness and population structure (GEMMA^31^ version 0.98.5). The GRM was estimated from SNPs with <1% missingness, a MAF >5%, info-score >0.95. PCAngsd^32^ was used to calculate per-site inbreeding coefficient (site-F) accounting for population structure. Sites with site-F > |0.05| and likelihood-ratio test P-value <1×10^−6^ were also excluded from the GRM estimation. Association tests assuming additive and recessive models were performed using a score test. Effect sizes and standard errors were estimated using a restricted maximum likelihood approach. Normalised protein expression (NPX) values were quantile transformed to a normal distribution using a rank-based inverse normal transformation performed separately for each sex. *P*-values are from analyses of transformed data and effect size estimates are from the same analyses and thus reported in standard deviations as β_SD_. In all analyses, sex, age, and cohort were included as covariates.

### Definition and refinement of significant loci

Primary pQTLs were first defined by distance-based clumping of ± 10Mb, reporting one representative SNP-protein association with the lowest P-value. For the proteins that are present in both the Cardiovascular II and Inflammation panels, we took pQTLs with the lowest P-value in either panel to avoid duplication. We reported the total number of identified significant associations using the conventional genome-wide threshold of 5×10^−8^.

Some SNP-protein pairs were genome-wide significant when tested in both additive and recessive models. To avoid double counting and to retain independence, if the same SNP-protein pair was genome-wide significant in both additive and recessive models, we reported the lowest p-value from the two, defined as ‘mostly additive’ if P_add_ < P_rec_ and ‘mostly recessive’ if P_rec_ < P_add_. SNPs within 1Mb significant for the same phenotype were also merged to the SNP with the lowest p-value, even if SNPs were detected with different models.

To test whether a SNP exhibited a ‘truly’ additive/recessive mode of inheritance, we tested whether the full model with full flexibility to the mode of inheritance (i.e. the alternative mode of inheritance is added as a covariate in the LMM) was significantly better than when using either an additive or recessive model, respectively. We subsequently defined the following categories for this test using an alpha threshold of 0.05: a) recessive/reject additive - where the full model is significantly better than the additive model, and the full model is not significantly better than the recessive model; b) additive/reject recessive - where the full model is not significantly better than the additive model, and the full model is significantly better than the recessive model; c) co-dominant/reject both - where the full model is significantly better than both the additive and recessive models, respectively.

To perform *cis/trans* classifications, we extracted the start/end information for all proteins by mapping UniProt IDs or gene names to Ensembl BioMart. Using these annotations, pQTLs were subsequently classified into four categories; for each protein, a *cis*-pQTL was defined when a lead SNP resided within 100 Kb upstream or downstream (± 100 Kb) of either the start or end of the gene encoding the associated protein, taking whichever was closer to the SNP; a *semi-cis*-pQTL was defined when a SNP resided within ± 100kb-5Mb of the gene encoding the associated protein; a *semi-trans*-pQTL was defined when a SNP resided within the same chromosome but >5Mb away; and a *trans*-pQTL was defined when a SNP was located in a different chromosome with respect to the corresponding protein-coding gene.

### Secondary associations and conditional analyses

To test for secondary signals, we performed conditional analyses where primary SNPs were used as covariates as a factor within both additive and recessive models. SNPs that were genome-wide significant when conditioning with any primary SNP within 10 Mb, were classified as a secondary association.

Similar to how we reported primary associations, secondary pQTL were classified as the lowest p-value from either the additive or recessive models, defined as ‘mostly additive’ if P_add_< P_rec_ and ‘mostly recessive’ if P_rec_ < P_add_. These secondary associations were then reported with their primary SNP.

### Proportion of variance explained by pQTLs

To ensure a fairer comparison when investigating effect sizes and proportion of variance explained (PVE) by pQTLs, we performed the same GWAS analysis described above using a random subset of 3,707 individuals from the UKB-PPP publication^14^. Of the 177 unique proteins assayed within this study, 176 were matched using UniProtIDs (data for VSIG2 was not available in the dataset described by Sun et al.^14^).

The following equation was used to estimate PVEs by pQTLs from GWAS summary statistics for each protein:

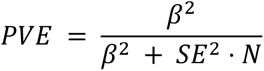

Where ß represents the effect size, SE represents the standard error of the effect size, and N represents the number of individuals assayed.

Comparisons were then made between our Greenlandic dataset (n=3,707), the UKB-PPP subset (n=3,707), and the UKB-PPP summary dataset (n=54,219; described in Supplementary Table 10 in Sun et al.^14^).

### Identification of novel pQTLs

To assess the novelty of the identified pQTLs in this study, we compared our results at genome-wide significance level to the results of 34 previously published pQTL studies, including the most recent UKB-PPP publication^14^. For all studies, we retained the pQTLs at the reported significance levels. We defined the novelty of locus-protein pairs if no variants residing within ± 10Mb of the primary pQTLs in this study had been reported in previous plasma/serum pQTL studies for the corresponding primary protein as in Thareja et al.^17^; described in Supplementary Table 4. Comparisons were done at protein level by matching the reported gene name from each study. Gene names were mapped based on Uniprot IDs, through Uniprot ID mapping in case of missing values.

In some cases, locus-protein pairs have been previously reported, yet the putative variant within this study could be novel. To identify such variants, we plotted SNP-protein effects from UK Biobank and the Greenlandic study together in Supplementary Figure 1 and for potentially novel pairs in Supplementary Figure 2.

### Analysis of known high-impact cardiometabolic-related variants

Summary data describing the associations between known high-impact Arctic-specific variants in *CPT1A* (rs80356779), *TBC1D4* (rs61736969), *HNF1A* (rs2135845768), *LDLR* (rs730882082), and *PCSK9* (rs4609471) on the abundance of 177 Olink proteins measured in this study were examined. False discovery rate (FDR) was calculated to account for multiple testing.

### Role of funding source

None of the funding agencies had any role in the study design or collection, analysis, or interpretation of the data.

## Results

### Genetic architecture of circulating plasma proteome

We analysed 177 proteins measured using the Olink Target Inflammation and Cardiovascular II panels (Supplementary Table 1) in three Greenlandic cohorts, totaling 3,707 participants from 19 different localities in Greenland (Supplementary Table 2). To identify pQTLs we used both an additive and a recessive model in a linear mixed model framework, which resulted in low genomic inflation for both the additive (mean λ= 0.985 ± 0.0246) and the recessive (λ= 0.989 ± 0.0250) model (Supplementary Figure 3). We identified a total of 251 genome-wide significant (*P*<5×10^−8^) primary associations between 225 genomic regions and 133 proteins (Figure 1, Supplementary Table 3), where primary associations means that all SNPs associated with the same protein are more than 10Mb apart. Of these primary pQTLs, 235 were classified as mostly additive and 16 as mostly recessive, however, for around one in four we can reject both the additive and the recessive model in favour of the full model (figure 1b). As expected, we found that the majority of primary SNPs were associated with a single protein, yet a large portion of proteins (56.4%) were associated with more than one SNP. In 34 previously published pQTL studies, we found that 181 of our identified locus-protein associations have been previously reported in a peer-reviewed article, and that these are in almost equal proportions in *cis* and *trans* (Figure 1f). Conversely, the vast majority of the remaining 70 (28%) locus-protein pairs were found to be novel (± 10Mb from the nearest variant previously reported to be associated to the same protein in a peer-reviewed article), in which the vast majority of these associations were in *trans* or semi-trans (96%; Figure 1f; Figure 1k, Supplementary Table 4). Interestingly, two of the three novel associations reported in *cis* that did not overlap with any of the examined published studies have lead SNPs that are very rare in European populations, yet were common in the Inuit ancestry component of the Greenlanders (chr16:28596376:T:C for ST1A1 - MAF=0.064 in Inuit and MAF=0.001 in Europeans; chr12:91158410:T:C for DCN - MAF=0.230 in Inuit and MAF<0.001 in Europeans; Supplementary Tables 3 & 4). Of the primary pQTLs, 93 (37%) were local-acting (85 *cis*, 8 *semi-cis*; Figure 1k; Methods) and 158 (63%) were distant-acting (14 *semi-trans* and 144 *trans*; Figure 1k; Methods). Of the 177 proteins assayed within this study, 51 unique proteins were identified as having both *cis* and *trans* pQTLs (defined as <±5Mb of the gene encoding the associated protein and >±5Mb of the gene encoding the associated protein or on a different chromosome, respectively). In addition to the 251 primary associations, conditional analyses further revealed 48 secondary pQTLs, of which, 47 were additive (36 *cis*, 3 *semi-cis;* 8 *trans*) and one recessive (in *cis*) (Supplementary Table 5).

**Figure 1.**
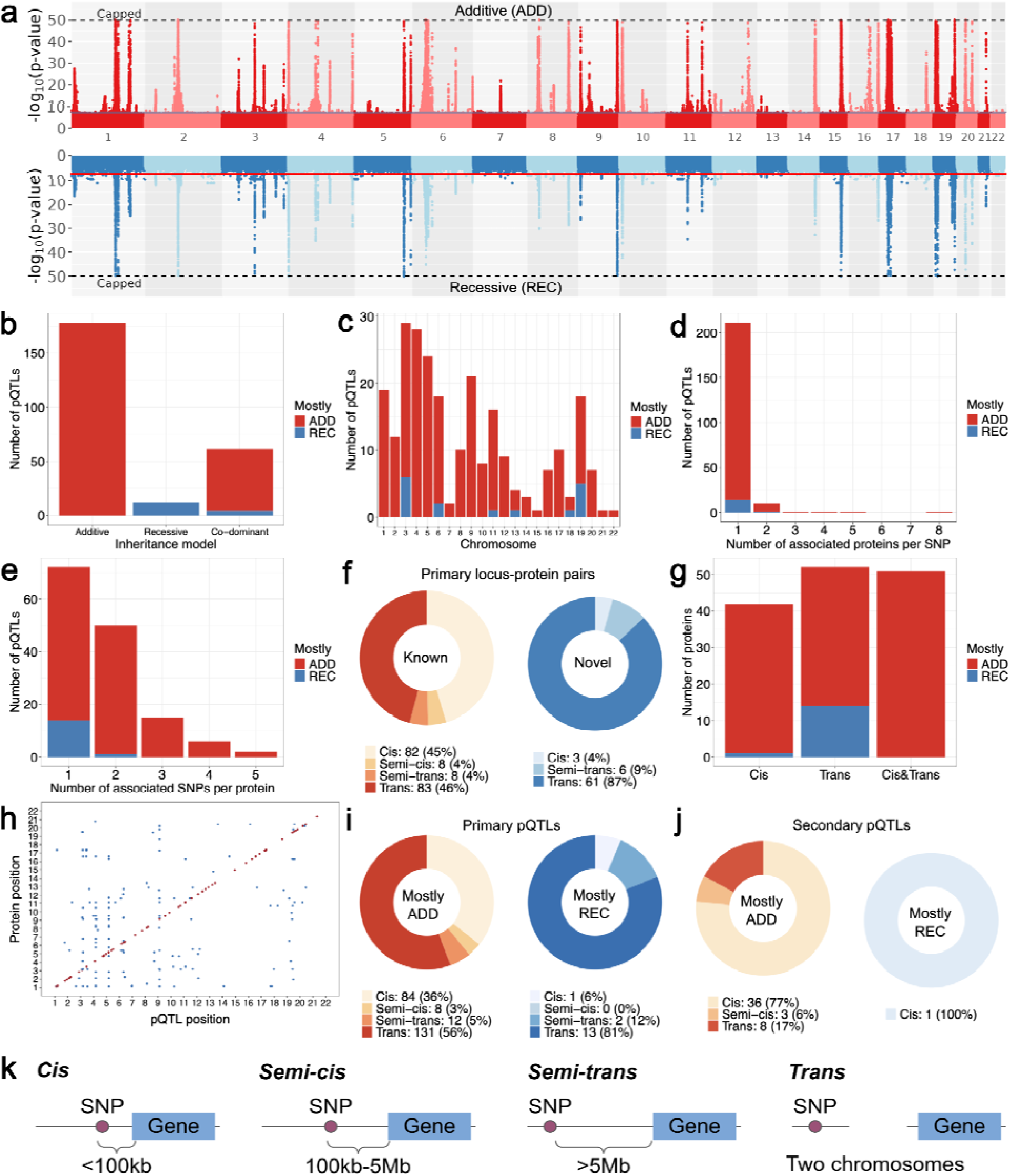
Genetic architecture of the targeted plasma proteome in the Greenlandic population. **a** Manhattan plot of p-values combined for all proteins under additive (red) and recessive (blue) models obtained using GEMMA. A genome-wide significance threshold of 5×10^-8^ is indicated by horizontal blue and red lines. P-values are capped at 1×10^−50^, as indicated by dotted black lines. **b** Primary pQTLs classified according to inheritance model where ‘additive’ and ‘recessive’ indicate pQTLs that are not rejected in favour of the full model while for ‘co-dominant’ both the additive and recessive models are rejected. Bar colours indicate the model in which the lowest p-value was reported, where red indicates a lower p-value when using an additive model (‘mostly additive’) and blue, a lower p-value when using a recessive model (‘mostly recessive’). **c** Number of primary pQTLs detected per chromosome. **d** Number of associated proteins per variant. **e** Number of primary pQTLs per protein. **f** Number of known and novel locus-protein associations in *cis*, *semi-cis*, *semi-trans* and *trans* (defined in k). Known indicates associations within ± 10Mb of any significant pQTL reported in previous studies (Methods). Novel indicates associations >± 10Mb of any significant pQTL reported in previous studies (Methods). **g** Number of proteins with pQTLs in *cis*, *trans* or both. *Trans* pQTLs here are defined as >5Mb from the start/end positions of the primary gene relevant for each Olink protein (a combination of *trans* and *semi-trans* as defined in k, for simplicity). **h** Dot plot indicating pQTL positions across the genome. *Trans* pQTLs are defined as >5Mb from the start/end positions of the primary gene relevant for each Olink protein, (a combination of *trans* and *semi-trans* as defined in k, for simplicity). Red - *cis* QTLs; Blue - *trans* pQTLs. **i** Number of primary pQTLs defined as *cis*, *semi-cis*, *semi-trans* and *trans* (defined in k). **j** Number of secondary pQTLs defined as *cis*, *semi-cis*, *semi-trans* and *trans* (defined in k). **k** Definitions of *cis*, *semi-cis*, *semi-trans* and *trans* pQTLs. *Cis* pQTLs are defined as SNP-protein pairs which are within 100kb of the start/end of the gene encoding the associated protein. *Semi-cis* pQTLs are defined as SNP-protein pairs which are between 100-kb-5Mb of the start/end of the gene encoding the associated protein. *Semi-trans* pQTLs are defined as SNP-protein pairs which are >5Mb of the start/end of the encoding gene, but on the same chromosome. *Trans* pQTLs are defined as SNP-protein pairs which are on different chromosomes from the gene encoding the associated protein.

### Common, high-impact pQTLs in Greenlanders

We next explored PVE for pQTLs detected in Greenlandic individuals and performed comparisons with Olink data from the UK Biobank^14^. The UK Biobank individuals tended to have a larger proportion of significant pQTLs with a lower PVE, and exhibited a lower number of high-impact pQTLs compared to the Greenlandic individuals (Figure 2a-b, Supplementary Table 6). In the Greenlandic individuals, several proteins, including IL-27, IgGFcRII-b, MCP-2 and IL-16 and Gal-9 exhibited PVEs >30% (Figure 2a). PQTLs related to the same proteins in UK Biobank exhibited PVEs <20% and were generally less common in allele frequency when compared to Greenlanders (Figure 2bi-iii). Two of these variants, rs1801274 associated with IgGFcRII-b, rs142653327 associated with Gal-9, are missense variants. Specifically, the association of IgGFcII-b rs1801274 G-allele has been shown to confer susceptibility to autoimmune diseases, including Kawasaki disease and ulcerative colitis^33^. Furthermore, rs142653327 is a novel pQTL variant for Gal-9, which is not present in Europe (Figure 2c). However, a secondary association for Gal-9 found in Greenland was also significant within UK Biobank (rs112305547; Figure 2c). Overall, these results suggest that Greenlanders exhibit more common and high-impact variants when compared to UK Biobank individuals, and that some of these variants may carry a functional impact.

**Figure 2.**
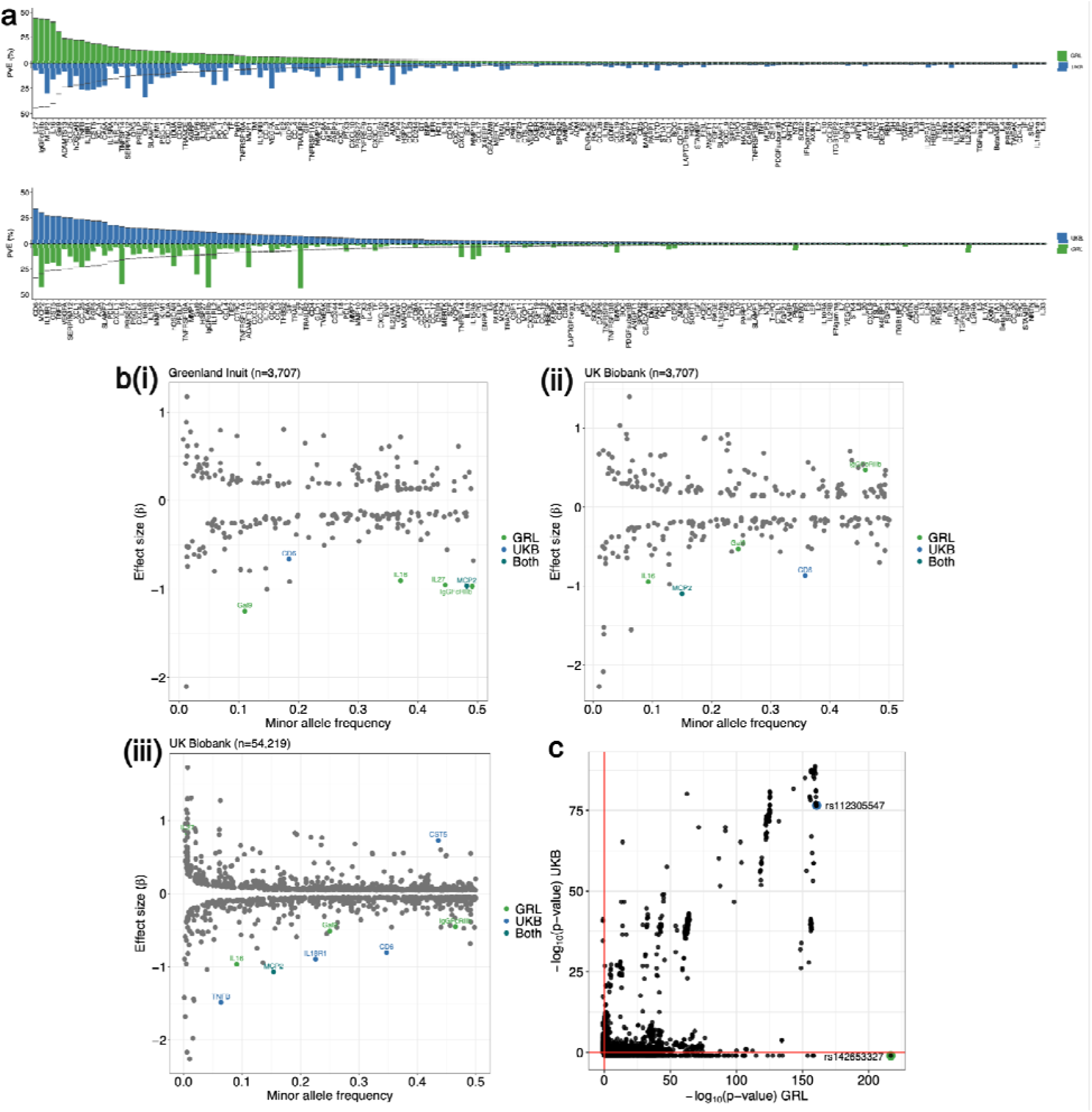
Proportion of variance explained by pQTLs in Greenlanders vs Europeans from UK Biobank. **a** Proportion of variance explained (PVE) for Greenlanders (GRL; green; n=3,707) and the UK Biobank cohort^14^ (UKB; blue; n=3,707) for 176 proteins common between both studies, ordered by highest PVEs in Greenlanders (top) and highest PVEs in UKB (bottom). Dashed lines indicate the equivalent PVE for each protein as reported in Greenlanders (top) and UKB (bottom), respectively. **b** PVE vs minor allele frequency (MAF) for genome-wide significant pQTLs detected in (i) Greenlandic individuals using GEMMA (n=3,707); (ii) a random subset of UKB individuals using GEMMA (n=3,707); and (iii) using summary statistics from Sun et al. (2024) (n=54,219). pQTLs with PVE >30% for either Greenlandic individuals, UKB individuals or both in (i) and (ii) are highlighted in green, blue and turquoise, respectively. PVE >30% for Greenlandic individuals, and PVE >15% for UKB individuals or meeting both criteria in (iii) are highlighted in green, blue and turquoise, respectively. **c** Comparisons of -log_10_ P values for variant-protein associations described by the Greenlanders dataset (n=3,707), and a subset of individuals from the UKB-PPP (n=3,707). Variant-protein associations tested using an additive model between Gal-9 and SNPs within 5Mb of rs142653327 are plotted. Red lines indicate -log_10_ P values of 0 for both datasets. Points at −1 indicate associations that were unable to be overlapped with the orthogonal dataset. Green circle - indicates the lead QTL in the Greenland cohort (rs142653327); Blue circle - indicates the lead secondary association reported in the Greenland cohort (rs112305547).

### Associations with known high-impact Arctic-specific variants

Previous studies have identified causal, high-impact variants related to population-specific effects for lipid metabolism, T2D, and CVD within Greenlanders, including *CPT1A*^34^, *TBC1D4*^35^, *HNF1A*^36^, *LDLR*^37^, and *PCSK9*^27^. As such, we investigated the protein expression for these known variants to gain a better understanding of potential mechanisms leading to these trait differences (Figure 3).

**Figure 3.**
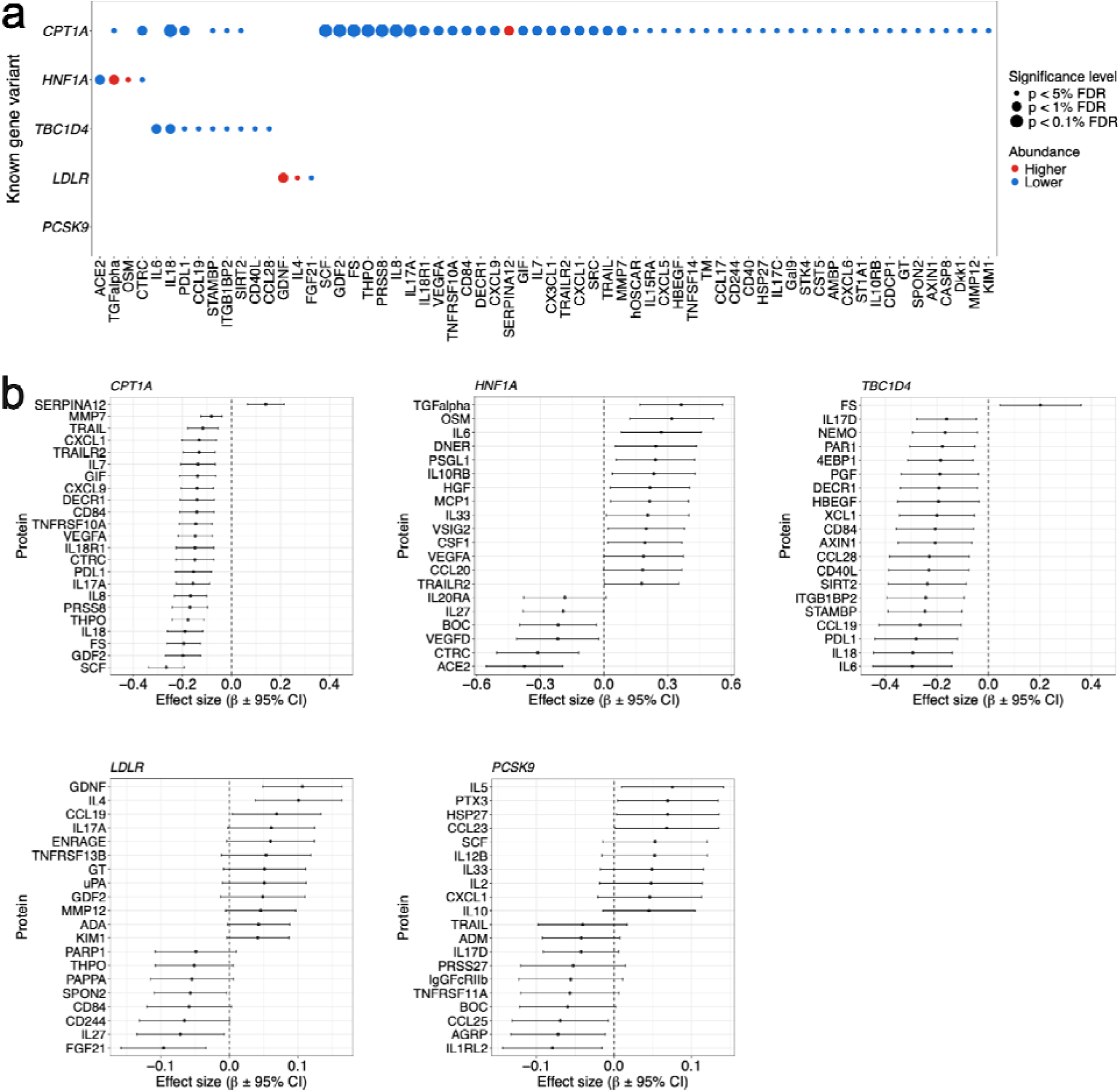
Known high-impact cardiometabolic-related Greenlandic variants. **a** Significant associations between known high-impact cardiometabolic-related Greenlandic variants - *CPT1A* (rs80356779), *HNF1A* (rs2135845768), *TBC1D4* (rs61736969), *LDLR* (rs730882082) and *PCSK9* (rs4609471) - and inflammatory and cardiovascular-related plasma proteins (FDR<0.05). Bubble sizes describe significance levels after multiple testing; small bubbles indicate pQTLs with FDR<0.05 and large bubbles FDR<0.01. Red - higher abundance; blue - lower abundance. **b** Forest plots for known high-impact variants within the same five genes and the top 20 proteins with the lowest FDR. The centre estimate and error bars on the x-axis describe the effect size (ß) ± 95% CI.

A known variant in *CPT1A* (rs80356779)^34^, previously found to modulate fatty acid composition, with large effect sizes on weight and height, herein was associated with lower plasma levels of 55 proteins at a significance level of FDR<0.05, including eight proteins significant at FDR<0.001. Stem cell factor, also termed kit ligand (SCF/KITLG) and follistatin (FS) represented the most significant proteins associated with this variant.

The known high-impact variant in *TBC1D4* (rs61736969)^35^, which causes insulin resistance in the skeletal muscle and explains a significant proportion of the variance in diabetes cases in Greenland, was associated with lower levels of nine plasma proteins after a 5% FDR correction (i.e. IL-6, IL-18, CCL-19, CCL-28, CD40L, ITGB1BP2, PD-L1, STAMBP and SIRT2).

A variant in the maturity onset diabetes of the young (MODY) gene, *HNF1A* (rs2135845768)^36^, which explains up to 2.5% of variance in diabetes in Greenland (together with *TBC1D4* explains up to 18% of diabetes cases), was associated with lower plasma levels of two proteins (angiotensin converting enzyme 2 [ACE2] and chymotrypsin-C [CTRC]) and higher levels of two proteins (oncostatin-M [OSM] and protransforming growth factor alpha [TGF-alpha]), after a 5% FDR correction.

A known variant in *LDLR* (rs730882082)^37^ which has a large impact on familial hypercholesterolemia (FH) in Greenland, was associated with lower plasma levels of one protein (fibroblast growth factor 21 [FGF-21]), and higher levels of two proteins (glial cell line-derived neurotrophic factor [GDNF] and interleukin-4 [IL-4]), after a 5% FDR correction.

The *PCSK9* loss-of-function variant (p.Arg46Leu; rs11591147) originally identified in European GWAS has led to PCSK9-inhibitors as a treatment for dyslipidemia^38^. We examined whether any proteins were associated with the recently identified *PCSK9* (rs4609471)^27^ variant in Greenland, but found no significant differences, after correction for multiple testing using FDR.

## Discussion

Here, we present a pQTL study in an isolated non-European population with 177 circulating inflammation- and cardiovascular-related proteins, measured using the Olink Proximity Extension Assay in 3,707 Greenlanders. We identified 251 genome-wide significant primary locus-protein pairs (93 *cis*, 158 *trans*), involving 225 different loci for 133 unique proteins. The 251 primary locus-protein pairs included many of the same locus-protein pairs found in prior studies performed in Europeans, with 181 (∼72.1%) reported previously in 34 peer-reviewed studies, but also revealed 70 locus-protein pair associations (3 *cis*, 6 *semi-trans*, 61 *trans*) that have not been reported previously. We identify 16 pQTLs which are purely recessive, 15 out of 16 are in *semi-trans/trans* (e.g. PRSS27, FABP2 and TF). This highlights the importance of conducting analyses in diverse populations, including alternative modes of inheritance.

Cross-ancestry differences have been highlighted by a previous mass-spectrometry based proteomics study conducted in a population of 2,958 Han Chinese participants, wherein proteins which are positively associated with BMI in European populations, showed opposite direction of effects in East Asians.^15^ An additional study found that up to 89% of pQTLs identified in European studies can be replicated in an Arab population of 2,935 individuals, when using the SOMAscan platform.^17^ So far, the pQTL studies performed in isolated populations are of European ancestry. An earlier study used a unique approach of whole-genome sequencing and three Olink panels (CVDI, CVDII, metabolism) across three isolated European cohorts (MANOLIS, Pomak, and ORCADES), identifying 5 rare variant pQTLs.^18^

To examine whether high-impact pQTLs were more common in the Greenlandic population, we performed an additional GWAS for a subset of UK Biobank individuals, matched by the sample size of our analysis for Greenlanders (n=3,707 in both populations) and using both additive and recessive models. The three pQTLs with the highest PVE in Greenlanders (IL-27, IgGFcRII-b, MCP-2) explained 44.3%, 43.7%, and 43.2%, compared to the top three pQTLs in the sample-size matched UK Biobank (CD6, MCP-2, IL-18R1) GWAS, which explained 33.9%, 30.0%, and 26.9%.

Certain genetic variants have undergone selection as an adaptation to the extreme conditions of the Arctic and the intake of specialised diets rich in proteins and polyunsaturated fatty acids (PUFAs). An example of this is the Arctic-specific variant in *CPT1A* (rs80356779), which is a known modulator of fatty acid composition (including decreased levels of circulating omega-6 and increased omega-3 PUFAs), which may affect the regulation of growth hormones, with large effects on height and weight^34^. Herein, we demonstrated that this variant has pleiotropic effects on plasma protein levels, with a general pattern of reduced inflammatory and cardiovascular-related protein biomarkers, which may be a result of the reduction in circulating PUFAs, primarily omega-6. *CPT1A* carriers exhibited significantly lower plasma levels of SCF, a protein that has been shown to play a central role in inflammation (cell survival, proliferation, and hematopoiesis) and is a suggested therapeutic target for inflammatory diseases^39^. *CPT1A* carriers also had significantly lower plasma levels of FS, a hepatokine which is elevated in patients with T2D and is associated with adipose tissue insulin resistance^40^.

The common loss-of-function variant in *TBC1D4*^35^ (rs61736969) responsible for muscle insulin resistance and explains up to 15.5% of variance in diabetes in Greenland, and in the present study was associated with lower expression of nine plasma proteins, most of which are interleukins (IL-6, IL-18), chemokines (CCL-19, CCL-28), other immune mediators (CD40L, PD-L1), integrins (ITGB1BP2), metalloproteases (STAMBP), and transferases (SIRT2). IL-6 is elevated with obesity, but on the other hand is also secreted as a myokine from muscles during exercise, exerting anti-inflammatory and anti-diabetogenic effects. Physical activity has been shown to reduce postprandial hyperglycemia in *TBC1D4* carriers^41^, and IL-6 levels may be increased as a result of exercise. Moreover, lower expression of ITGB1BP2 (integrin beta-1-binding protein 2) was also observed in *TBC1D4* carriers, a protein that plays an essential role in the maturation and organisation of muscle cells^42^, which could be driven by the impaired insulin resistance in skeletal muscle. It remains unclear whether *TBC1D4* carriers are at an increased risk of cardiovascular disease^43^. In the present study, we see elevations in several cardiovascular-disease related biomarkers, including ITGB1BP2^42^, STAMBP (linked to congenital malformations of the heart)^44^ and SIRT2 (functions in inhibiting pathological cardiac hypertrophy)^45^.

The known Inuit-specific variant in the MODY gene *HNF1A*^36^ (rs2135845768), explaining up to 2.5% of variance in diabetes in Greenland, was associated with altered expression of four (ACE2, CTRC, OSM, TGF-alpha) proteins in the present study. Plasma abundance of ACE2 was significantly lower in *HNF1A* carriers. *HNF1A* is known to induce ACE2 expression in the pancreatic islet cells. ACE2 plays an important role in maintaining glycemia and β-cell function, and has been suggested as a potential therapeutic target for treatment of diabetes^46^.

A known Arctic-specific variant in *LDLR*^37^ (rs730882082) which poses a high risk of FH in nearly 30% of Greenlanders, associated with altered expression of three plasma proteins (FGF-21, GDNF, and IL-4) in the present study. Specifically, FGF-21 levels were lower in carriers of the *LDLR* variant, a protein responsible for fatty acid oxidation and regulation of systemic glucose homeostasis and insulin sensitivity. FGF-21 analogs are being developed for their beneficial effects on modulate lipid and lipoprotein levels, and therefore could potentially serve as treatment for patients with FH^47^. Clinical trials for FGF-21 analogs are also ongoing for metabolic-dysfunction-associated steatotic liver disease.

### Limitations

Our study has several limitations. Firstly, our pQTL analysis is restricted to the 177 proteins measured by two Olink panels. An untargeted approach would allow for identification of more and potentially novel pQTLs, yet targeted approaches allow for the study of biologically relevant protein biomarkers. Inclusion of orthogonal methods such as mass spectrometry-based proteomics, would resolve the possibility of epitope effects, yet this kind of validation is difficult to perform at scale. Tissue- and cell-type specific pQTL studies will be valuable in future studies to understand differences in genetic signals across tissues. We did not examine ratios between protein levels which may identify novel associations, when multiple proteins are associated with the same variant^48^. Furthermore, the raw abundance of proteins does not take into consideration protein-protein interactions, mapping the interactome can lead to additional biological insights^49^.

### Strengths

Identification of pQTLs aid in the understanding of mechanisms of complex disease and can identify promising targets for pharmacological interventions. The majority of all pQTL studies performed to date have been conducted in European populations, limiting the generalizability of findings across populations. Although an attempt was made to identify pQTLs in non-European ancestry in the UKB-PPP (953 with South Asian and 1,513 with African ancestry), the sample size is smaller than the present study^50^. Here, we explore pQTLs in a historically isolated Greenlandic population, identify associations related to alternative modes of inheritance, and highlight the need to study diverse populations, through comparisons to large datasets generated in the UK Biobank, primarily of European ancestry. The challenge in genetic research in indigenous populations is that it raises complex psychological, ethical, social, and political issues. These issues have been raised in a series of user surveys on the use of genetics in metabolic research. Generally, the communities fully support the research with a specific request to investigate the significance of traditional and modern Inuit lifestyle, in order to understand how advising individuals regarding diet and lifestyle, can be linked in a culturally appropriate way, to better inform health effects of genetic profiles.

## Conclusions

The pQTLs identified and analysed in this study provide a resource to unravel the genetic architecture of the targeted plasma proteome in the Greenlandic population. There was a larger proportional variance explained in Greenlanders when compared to European ancestries, which has relevant implications for disease outcomes. Variants with large population impact on lipid metabolism (*CPT1A*) and risk of cardiometabolic disease in Greenland (*TBC1D4*, *HNF1A*, and *LDLR*), show distinct patterns with inflammatory cytokines and cardiovascular disease biomarkers, pointing towards potential therapeutic targets.

## Supporting information

Description of supplemental figures

SuppFig1

SuppFig2

SuppFig3

## Data Availability

Summary data from the study may be available by reasonable request to the authors.

## Contributors

● Conceptualization: SES, RFB, MKA, NG, MEJ, IM, AA, TH
● Data curation: SES, MKA, ACBT, NG, MEJ, IM, AA, TH
● Formal analysis: SES, RFB, FFS, SH, LL
● Funding acquisition: PB, CVLL, NG, MEJ, IM, AA, TH
● Investigation: SES, RFB, FFS, PB, CVLL, NG, MEJ, IM, AA, TH
● Methodology: SES, RFB, FFS, NG, MEJ, IM, AA, TH
● Project administration: SES, RFB, MKA, PB, CVLL, NG, MEJ, IM, AA, TH
● Resources: PB, CVLL, NG, MEJ, IM, AA, TH
● Software: AA
● Supervision: NG, IM, AA, TH
● User surveys: MEJ
● Visualisation: SES, RFB, FFS, IM, AA, TH
● Writing - original draft: SES, RFB, IM, AA, TH
● Writing - review & editing: MKA, FFS, SH, ACBT, LL, PB, CVLL, NG, MEJ

## Data sharing statement

Summary data from the study may be available by reasonable request to the authors.

## Declaration of interests

The population surveys of Greenland were funded by the Department of Health, Greenland, The Novo Nordisk Foundation (NNF17OC0028136 & NNF17SH0027192), The Independent Research Fund Denmark, and Karen Elise Jensen Foundation. The Novo Nordisk Foundation Center for Basic Metabolic Research is an independent research centre at the University of Copenhagen partially funded by an unrestricted donation from the Novo Nordisk Foundation (NNF18CC0034900). SES received funding from The Novo Nordisk Foundation, Copenhagen Bioscience PhD Program (grant number: NNF18CC0033668). RFB and IM are supported by a Villum Young Investigator grant (VIL19114), awarded to IM. RFB, SH, LL and AA are funded by the Novo Nordisk Foundation (NNF20OC0061343). AA is supported by the Independent Research Fund Denmark (8021-00360B). None of the funding agencies had any role in study design or collection, analysis, or interpretation of data. NG is currently employed at Novo Nordisk. All other authors have no further conflicts of interest to declare.

## Acknowledgements

We would like to acknowledge all the participants and the staff of the Greenland Population Health Surveys (Befolkningsundersøgelsen i Grønland). We would also like to thank Annemette Forman and Tina Hvidtfeldt Lorentzen from the Novo Nordisk Foundation Center for Basic Metabolic Research, University of Copenhagen for their technical support.

## Appendix A. Supplementary Data

## Appendix B. Supplementary Figures

## Notes

### Competing Interest Statement

The authors have declared no competing interest.

### Funding Statement

Novo Nordisk Foundation, Independent Research Fund Denmark, and Karen Elise Jensen Foundation

### Author Declarations

The study has received ethical approval from the Science Ethics Committee in Greenland. The study was conducted in accordance with the Declaration of Helsinki, second revision. All participants gave written informed consent.

### Summary of Updates

Figure 1 and 2 revised.

