## Supplementary material for "Genetic regulation of the plasma proteome and its link to cardiometabolic disease in Greenlandic Inuit": Description of supplemental figures

**Description of Supplementary Figures**

**Supplementary Figure 1.** Comparisons of -log_10_ P values for variant-protein associations described by the Greenland Inuit dataset (n=3,707), and a subset of individuals from the UKB-PPP (n=3,707)^1^. Associations tested using additive and recessive models within 5Mb of each genome-wide significant (P<5e^-8^) Inuit pQTL are plotted here (additive - left and recessive - right panels for each pQTL, respectively). Values for 248/251 pQTLs detected within this study are depicted (3 pQTLs were detected for VSIG2; protein expression data was unavailable for the UKB-PPP cohort for this protein). Points at -1 indicate associations that were unable to be overlapped with the orthogonal set. Red lines indicate -log_10_ P values of 0 for both datasets.

**Supplementary Figure 2.** Comparisons of -log_10_ P values for 79 ‘novel’ pQTLs (as defined by ± 1Mb flanking regions from the sentinel variant), within the Greenland Inuit dataset (n=3,707) and a subset of individuals from the UKB-PPP (n=3,707)^1^. Associations tested using additive and recessive models within 5Mb of each novel Inuit pQTL are plotted here (additive - left and recessive - right panels for each pQTL, respectively). Values for 76/79 pQTLs detected within this study are depicted (3 pQTLs were detected for VSIG2; protein expression data was unavailable for the UKB-PPP cohort for this protein). Red lines indicate -log_10_ P values of 0 for both datasets. Points at -1 indicate associations that were unable to be overlapped with the orthogonal dataset.

**Supplementary Figure 3.** QQ-plots for all 177 tested proteins for both additive (left) and recessive (right) models, with reported mean lambda values. Observed P values are capped 1x10^-30^.
