## Supplementary figures and images for "Genetic regulation of the plasma proteome and its link to cardiometabolic disease in Greenlandic Inuit"

### SuppFig1

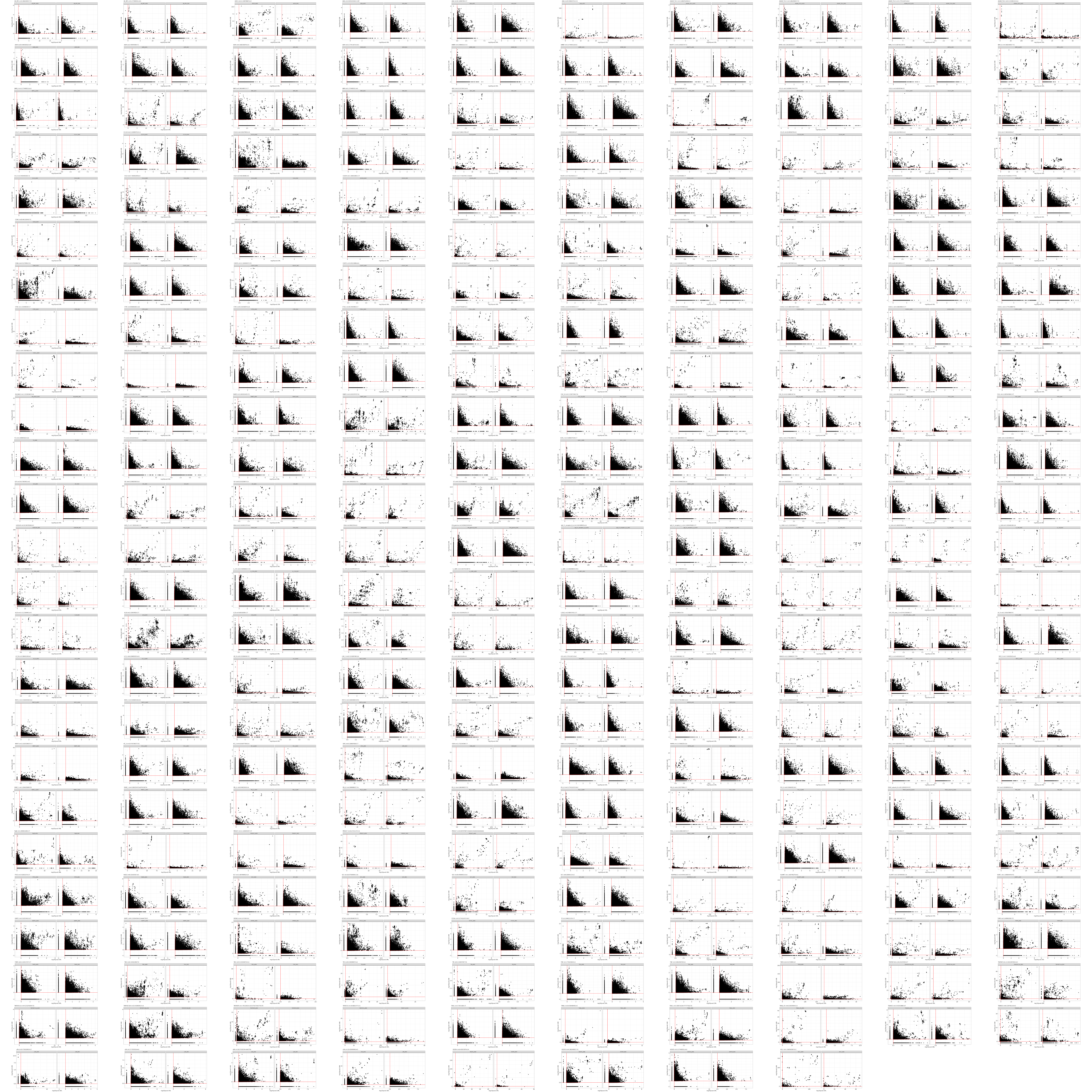

### SuppFig2

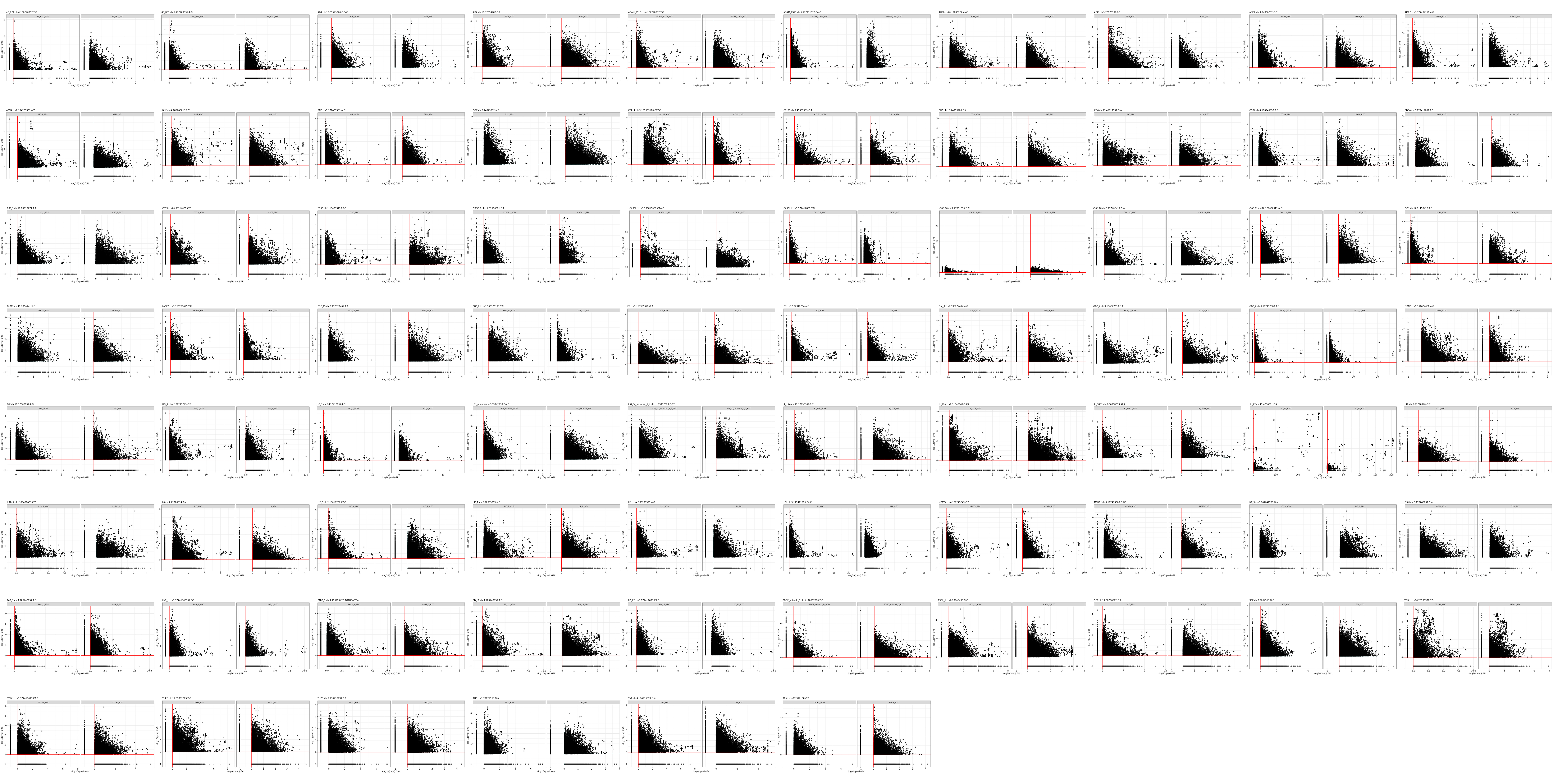

### SuppFig3

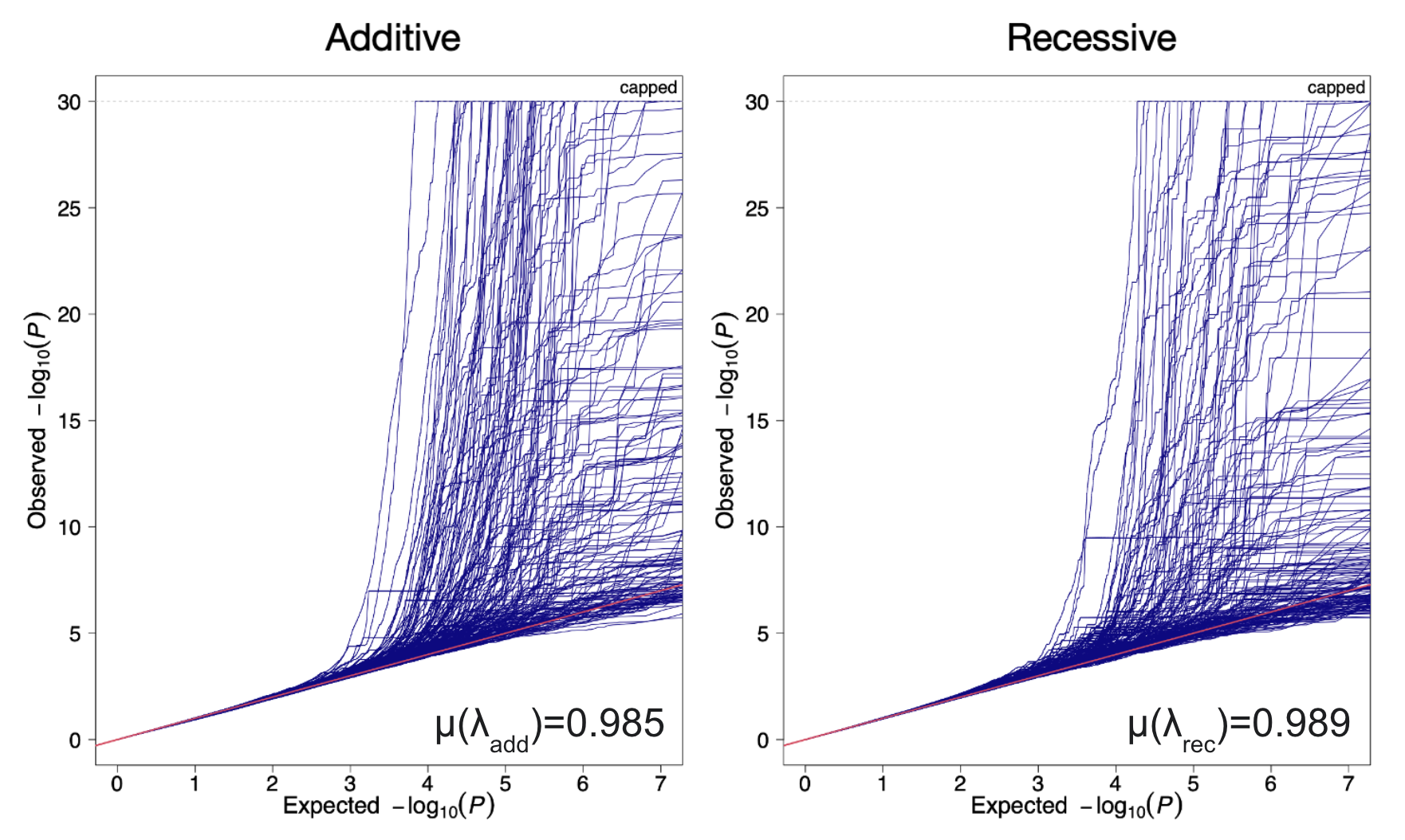
